# Defining an Ageing-Related Pathology, Disease or Syndrome: International Consensus Statement

**DOI:** 10.1101/2024.09.02.24312951

**Authors:** Emma Short, Ian M. Adcock, Bilal Al-Sarireh, Ann Ager, Ramzi Ajjan, Naveed Akbar, Michael A. Akeroyd, Ghada Alsaleh, Ghada Al-Sharbatee, Kambiz Alavian, Winfried Amoaku, Julie Andersen, Chrystalina Antoniades, Mark J. Arends, Sue Astley, Denize Atan, Richard Attanoos, Johannes Attems, Steve Bain, Konstantinos Balaskas, Gabriel Balmus, Manohar Bance, Thomas M. Barber, Ajoy Bardhan, Karen Barker, Peter Barnes, Gemma Basatemur, Adrian Bateman, Moises Evandro Bauer, Christopher Bellamy, Edwin van Beek, Ilaria Bellantuono, Emyr Benbow, Sunil Bhandari, Rahul Bhatnagar, Philip Bloom, Dawn Bowdish, Melissa Bowerman, Melanie Burke, Roxana Carare, Emma Victoria Carrington, Jorge Iván Castillo-Quan, Peter Clegg, James Cole, Carlo Cota, Paul Chazot, Christopher Chen, Ying Cheong, Gary Christopher, George Church, David Clancy, Paul Cool, Francesco Del Galdo, Mayank Dalakoti, Soumit Dasgupta, Colleen Deane, Devesh Dhasmana, Stefan Dojcinov, Monia Di Prete, Huaidong Du, Niharika A Duggal, Toby Ellmers, Costanza Emanueli, Mark Emberton, Jorge D. Erusalimsky, Laurence Feldmeyer, Alexander Fleming, Karen Forbes, Thomas C. Foster, Daniela Frasca, Ian Frayling, Daniel Freedman, Tamas Fülöp, Georgina Ellison-Hughes, Gus Gazzard, Christopher George, Jesus Gil, Richard Glassock, Rob Goldin, John Green, Robyn Guymer, Hasan Haboubi, Lorna Harries, Simon Hart, Douglas Hartley, Sebri Hasaballa, Christin Henein, Maggie Helliwell, Emily Henderson, Rakesh Heer, Kristofer Holte, Iskander Idris, David Isenburg, Juulia Jylhävä, Ahmed Iqbal, Simon W. Jones, Rajesh Kalaria, Venkateswarlu Kanamarlapudi, Werner Kempf, Alexandra J. Kermack, Jemma Kerns, Albert Koulman, Adnan H. Khan, James Kinross, Katarina Klaucane, Yamini Krishna, Harinderjit Singh Gill, Edward Lakatta, Ezio Laconi, Alpar Lazar, Christiaan Leeuwenburgh, Samantha Leung, Xuan Li, Ian van der Linde, Luísa V. Lopes, Antonello Lorenzini, Andrew Lotery, Pedro Machado, Sarah Mackie, Paolo Madeddu, Andrea Maier, Krishna Mukkanna, Pinelopi Manousou, Oonagh Markey, Claudio Mauro, Barry McDonnell, Reinhold J. Medina, Soma Meran, Claudia Metzler-Baddeley, Ignor Meglinksi, Neta Milman, Christina Mitteldorf, Ruth Montgomery, Andrew Conway Morris, Beda Mühleisen, Abhik Mukherkee, Andrew Murray, Scott Nelson, Anna Nicolaou, Alexander Nirenberg, Simon Noble, Lisa S. Nolan, Meritxell Nus, Canh Van On, Victor Osei-Lah, Mandy Peffers, Antony Palmer, Donald Palmer, Laura Palmer, William Parry-Smith, Graham Pawelec, Shahaf Peleg, Ranmith Perera, Andrew Pitsillides, Christopher J. Plack, Franze Progatzsky, Sonja Pyott, Kaukab Rajput, Sameera Rashid, J. Arjuna Ratnayaka, Sudhira A.B. Ratnayake, Manuel Rodriguez-Justo, Arianna Carolina Rosa, Andrew Rule, Gareth J. Sanger, Ian Sayers, Andrew Saykin, Dinesh Selvarajah, Jaswinder Sethi, Cathy Shanahan, Shai Shen-Orr, Carl Sheridan, Paul Shiels, Kastytis Sidlauskas, Sobha Sivaprasad, Judith Sluimer, Gary Small, Peter Smith, Rebecca Smith, Sarah Snelling, Ioakim Spyridopoulos, Ramasamy Srinivasa Raghavan, David Steel, Karen P Steel, Claire Stewart, Keeron Stone, Selvarani Subbarayan, Mark Sussman, Jonas Svensson, Vyshnavi Tadanki, Ai Lyn Tan, Rudolph E. Tanzi, Amanda Tatler, Adriana A. S. Tavares, Tengku Amatullah Madeehah Tengku Mohd, Ana Tiganescu, James Timmons, Jeremy Tree, Drupad Trivedi, Emmanuel A. Tsochatzis, Dialechti Tsimpida, Elisabeth J Vinke, Anna Whittaker, Neeru A. Vallabh, Kristin Veighey, Zoe C. Venables, Venkat Reddy, Meike W. Vernooij, Chris Verschoor, Manlio Vinciguerra, Vesna Vukanovic, Vladyslav Vyazovskiy, James Walker, Richard Wakefield, Adam J. Watkins, Anthony Webster, Caroline Weight, Birgit Weinberger, Susan L. Whitney, Rosalind Willis, Jacek M. Witkowski, Leonard L.L. Yeo, Tham Yhi Chung, Emma Yu, Michael Zemel, Stuart R.G. Calimport, Barry L. Bentley

**Affiliations:** Cardiff Metropolitan University, UK; Swansea Bay University Health Board, UK; Imperial College, UK; Cardiff University, UK; University of Leeds, UK; Oxford University, UK; University of Nottingham, UK; Wessex Deanery, UK; Buck Institute for Research on Ageing, California, USA; University of Edinburgh, UK; University of Manchester, UK; University of Bristol, UK; Cardiff and Vale University Health Board, UK; Newcastle University, UK; Swansea University, UK; Moorfields Eye Hospital, UK; University College London, UK; University of Cambridge, UK; Transylvanian Institute of Neuroscience, Cluj-Napoca, Romania; Warwick University, UK; University Hospitals Coventry and Warwickshire NHS Trust, UK; University of Birmingham, UK; University of Southampton, UK; Pontifical Catholic University of Rio Grande do Sul, Brazil; University of Sheffield, UK; Hull University Teaching Hospitals NHS Trust, UK; Hull York Medical School, UK; North Bristol NHS Trust; University of Southern Denmark, Denmark; McMaster University, Canada; Keele University, UK; Robert Jones and Agnes Hunt Orthopaedic Hospital, Oswestry, UK; Harvard Medical School, USA; University of Liverpool, UK; San Gallicano Dermatological Institute, Rome; Durham University, UK; National University of Singapore, Singapore; Lancaster University, UK; National University Heart Centre, Singapore; Alder Hey Children’s Hospital, UK; St Andrew’s University, UK; San Gallicano Dermatological Institute, Rome, UK; Bern University Hospital, Switzerland; Kinexum, USA; University of Florida, USA; University of Miami Miller School of Medicine, USA; Surrey and Sussex NHS Trust, UK; University of Sherbrooke, Canada; Kings College London, UK; NIHR Biomedical Research Centre, UK; MRC Laboratory of Medical Sciences, UK; Geffen School of Medicine at UCLA, USA; Centre for Eye Research Australia, University of Melbourne, Australia; University of Exeter, UK; National Institute for Health Research Biomedical Research Centre for Ophthalmology, UK; Royal Victoria Hospital, Belfast, UK; Karolinska Institutet, Stockholm, Sweden; University of Tampere, Tampere, Finland; University Hospital Zurich, Switzerland; University of Surrey, UK; The University of Iowa, Iowa City, Iowa, USA; St Thomas’s Hospital, London, UK; Liverpool University Hospitals NHS Foundation Trust, UK; University of Bath, UK; National Institute on Aging, NIH, USA; Università di Cagliari, Italy; University of East Anglia, UK; Lagan Valley Hospital, UK; Anglia Ruskin University, UK; Universidade de Lisboa, Portugal; Leeds Teaching Hospitals NHS Trust, UK; University of Amsterdam, Netherlands; Imperial College Healthcare NHS Trust, UK; Loughborough University, UK; Aston University, UK; Technion Israel Institute of Technology, Israel; University Medical Center Goettingen, Germany; Yale School of Medicine, USA; University of Basel, Switzerland; University of Glasgow, UK; Australasian College of Cutaneous Oncology, Victoria, Australia; Portsmouth Hospitals University NHS Trust, UK; Royal Veterinary College, UK; University of Tübingen, Germany; Health Sciences North Research Institute, ON, Canada; FBN Dummerstorf, Germany; Guy’s and St Thomas’ NHS Foundation Trust, UK; Great Ormond Street Hospital, UK; The Christie NHS Foundation, UK; University of Turin, Italy; Mayo Clinic, USA; Queen Mary University of London, UK; Indiana University School of Medicine, USA; Maastricht University Medical Center, Maastricht, The Netherlands; RWTH Aachen University, Medical Faculty, Aachen, Germany; Hackensack Meridian School of Medicine, USA; Royal Surrey NHS Foundation Trust, UK; Sunderland Eye Infirmary, UK; Liverpool John Moores University, UK; National Cardiovascular Research network, Wales, UK; University of Aberdeen, UK; San Diego State University, USA; Massachusetts General Hospital, USA; Universiti Sains Islam, Malaysia; Erasmus MC University Medical Centre Rotterdam, Netherlands; University of Stirling, UK; Norfolk and Norwich University Hospital, UK; Norwich Medical School, UK; Medical University of Varna, Varna, Bulgaria; Universität Innsbruck, Institute for Biomedical Ageing Research, Austria; University of Pittsburgh, USA; Medical University of Gdańsk, Poland; University of Tennessee, USA; Shriners Children’s, Boston, MA, USA; University of Bologna, Italy

## Abstract

**Background:** Around the world, individuals are living longer, but an increased average lifespan does not always equate to an increased healthspan. With advancing age, the increased prevalence of ageing-related diseases can have a significant impact on health status, functional capacity, and quality of life. It is therefore vital to develop comprehensive classification and staging systems for ageing-related pathologies, diseases and syndromes. This will allow societies to better identify, quantify, understand, and meet the healthcare, workforce, wellbeing, and socioeconomic needs of ageing populations, while supporting the development and utilisation of interventions to prevent or to slow, halt or reverse the progression of ageing-related pathologies.

**Methods:** The foundation for developing such classification and staging systems is to define the scope of what constitutes an ageing-related pathology, disease or syndrome. To this end, a consensus meeting was hosted by the International Consortium to Classify Ageing-Related Pathologies (ICCARP), on February 19^th^, 2024, in Cardiff, UK, and was attended by 150 recognised experts. Discussions and voting were centred on provisional criteria that had been distributed prior to the meeting. The participants debated and voted on these. Each criterion required a consensus agreement of ≥70% for approval.

**Results:** The accepted criteria for an ageing-related pathology, disease or syndrome were:

1. Develops and/or progresses with increasing chronological age.
2. Should be associated with, or contribute to, functional decline, or an increased susceptibility to functional decline.
3. Evidenced by studies in humans.

**Conclusions:** Criteria for an ageing-related pathology, disease or syndrome have been agreed by an international consortium of subject experts. These criteria will now be used by the ICCARP for the classification and ultimately staging of ageing-related pathologies, diseases and syndromes.

## Introduction

The World Health Organization has reported that around the world, people are living longer, and that every country is experiencing growth in both the number and the proportion of older people in the population [1]. Despite increasing average longevity, evidence suggests that the proportion of years lived in good health has remained broadly constant, which implies that some of the additional years of life are spent in poorer health [1] and many individuals live with multimorbidity. Ageing is characterised *inter alia* by the time-related progressive accumulation of damage, which can occur at molecular, cellular, tissue, organ, and system levels. This can have a detrimental effect on an individual’s intrinsic capacity, and can impact physiological, cognitive, psychological, and social functioning, and/ or socioeconomic status and productivity levels.

Over recent decades, research has led to a significant increase in the understanding of the biological features of ageing, but this has not yet been translated into clinically relevant classification and staging systems for ageing-related pathologies. In the International Classification of Diseases, 11^th^ revision (ICD-11) [2], there is a causality code related to ageing (XT9T) [2, 3, 4] to classify entities *‘caused by biological processes which persistently lead to the loss of organism’s adaptation and progress in older ages’* [2] and a code under ‘General symptoms, signs or clinical findings’ (MG2A) for *‘ageing associated decline in intrinsic capacity*.*’* [2] Furthermore, there are several entities that are described as being associated with increasing age, for example photoageing of the skin, intrinsic ageing of the skin or hearing loss. However, the existing approach to ageing-related pathologies is superficial and non-standardised.

In 2019, Calimport *et al*. called for the systematic and comprehensive classification and staging of ageing-related pathologies at the metabolic, tissue, organ, and systemic levels [5]. It was recommended that such a classification system should be adopted by the ICD to guide policy and practice as well as to enable appropriate clinical guidance, systems, resources, and infrastructure [5]. However, progress in developing such classification and staging systems has been slow, and there is an urgent need for accelerated efforts to identify, characterise, name and classify ageing-related pathologies, diseases and syndromes.

To this end, the International Consortium for the Classification of Ageing-Related Pathologies (ICCARP) was established in 2023, comprising 16 international working groups, initially to develop the classification systems. The ICCARP is led by a research team at Cardiff Metropolitan University. Fourteen working groups have been structured on a system-specific basis: audiovestibular; breast; cardiovascular; dermatology; endocrine and metabolic; gastrointestinal, pancreatic, hepatobiliary; gynaecology; immunology; musculoskeletal; nephrology; neurology; ophthalmology; respiratory and urology. In addition, there are scientific advisory and standardisation groups. Overall, the working groups comprise around 300 clinicians, research scientists and allied health professionals who are recognised subject experts in their fields.

## Methods

The hybrid International Consensus Meeting to Define an Ageing-Related Pathology, Disease or Syndrome was hosted in Cardiff, UK, on February 19^th^, 2024. Before the meeting, the primary research team (Dr Emma Short, Dr Barry Bentley and Dr Stuart Calimport, Cardiff Metropolitan University) had developed five potential criteria to define an ageing-related pathology, disease or syndrome:

1. Must develop/ progress with increasing age.
2. Must cause functional decline.
3. Must predict mortality.
4. Evidenced by studies in humans.
5. Mendelian disorders are excluded. These criteria were distributed to all ICCARP working group members one month before the meeting, for comments and feedback. Based on the feedback received during this initial consultation period, a sixth criterion was added:
6. Should not be primarily accounted for by an extrinsic carcinogen/ environmental toxin/ infectious agent/ injury.

150 working group members attended the meeting, representing 65 different institutions from 15 countries. Most individuals participated virtually (93%). Each criterion was debated sequentially, and where relevant, refined wording was suggested. At the end of each discussion, participants were invited to vote as to whether they agreed with the criterion. Voting options were: “Yes”, “Yes with reservations”, “No”, and “Abstain”. Reservations raised were subsequently discussed, and participants were re-polled where modified wording had been proposed. Voting was performed through a Teams anonymous online poll for the virtual attendees and through the raising of hands for in-person attendees. Each criterion required a consensus agreement of ≥70% for approval, consisting of “Yes” or “Yes with reservations”.

## Results

The criteria for defining an ageing-related pathology, disease or syndrome that were accepted were: (1) Develops and/ or progresses with increasing chronological age, (2) Should be associated with, or contribute to, functional decline, or an increased susceptibility to functional decline, and (3) Evidenced by studies in humans.

### 1. Develops and/ or progresses with increasing chronological age

The first criterion was agreed by 97% of the consortium.

There was considerable discussion surrounding whether the criterion should include a specific age-related threshold. For example, it was suggested that the criterion should explicitly state that it was referring to pathologies of adulthood. However, it was acknowledged that ‘adulthood’ potentially has different definitions, including a chronological age of 18 years *or* when an individual reaches skeletal maturity *or* at the end of puberty, and it was highlighted that the maturation of different body systems can occur at different chronological ages. Furthermore, it was recognised that several chronic diseases traditionally thought of as ‘older age-related diseases’, for example, type 2 diabetes mellitus, are now being observed in younger populations [6] and that some ageing-related pathologies can even be present from the time of conception. It is vital that pathologies can be identified at very early stages for purposes of reversal or prevention of progression.

The conclusion was to avoid an arbitrary cut-off point, since some phenomena attributed to ageing can be identified in chronologically young individuals. It would not be appropriate to exclude such individuals from clinically relevant classification and staging systems.

### 2. Should be associated with, or contribute to, functional decline, or an increased susceptibility to functional decline

There was 99% agreement with the inclusion of criterion 2, with the understanding that functional decline refers to a decrement in physiological, physical, cognitive or socioeconomic functioning, but the working groups would define the specific details on a system-specific basis.

The original suggestion, ‘must cause functional decline’ was reformulated based on several points raised by the working group members. ‘Must’ was considered too restrictive, so was amended to ‘should’. It was reinforced that ageing-related pathologies do not always *cause* functional decline, but rather, some play a *contributory* role, alongside other contributory factors, and some do not directly result in functional decline but are *associated* with functional decline or have a bi-directional relationship. Furthermore, it was acknowledged that some ageing-related pathologies might be silent, especially if there is functional reserve within the system, and will only become apparent if there is an additional insult or an additional pathology develops. For example, clonal haematopoiesis can be clinically silent [7], but it can manifest as a patient becoming systemically unwell because of an infection (the additional insult), even if it does not develop into myelodysplasia or leukaemia. To recognise this, the criterion was reworded to include ‘increased susceptibility to functional decline’.

Whilst some participants suggested that ageing-related functional decline should be defined as ‘decline that is irreversible’, this was rejected by the majority of the consortium, who agreed that functional decline may be transient or chronic but *need not* be permanent, for example it may be reversed following an intervention. Whilst some functional decline may be irreversible at the current time, this is not always true, and it is hoped that some declines could be halted or reversed once mechanistic causes are identified.

### 3. Evidenced by studies in humans

92% of the consortium agreed with criterion 3.

It is recognised that whilst data from animal studies may provide supporting evidence and can help understand disease mechanisms and develop treatments, it is imperative that there is significant evidence from studies in humans. The ICD is a classification system for humans only and as such, relies on evidence from studies in humans.

**Figure 1:**
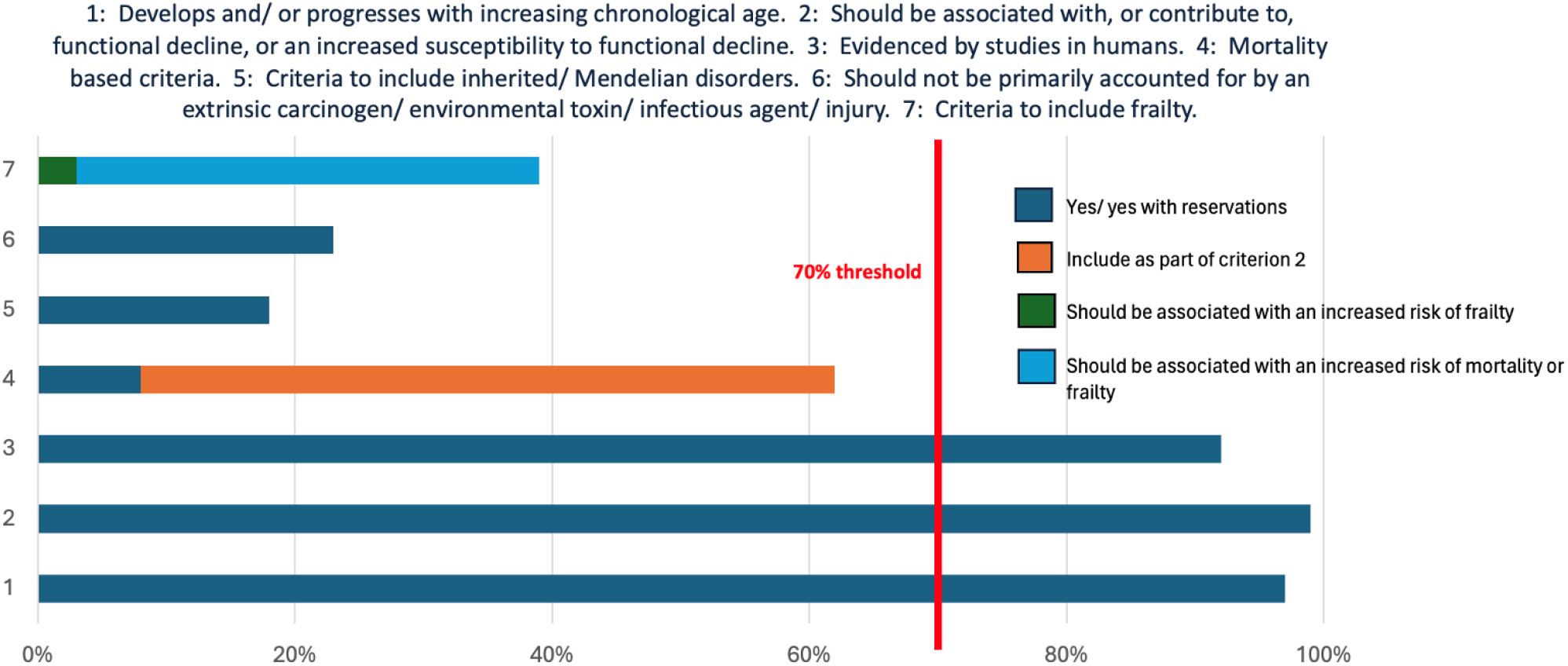
% agreement with criteria

## Rejected criteria and further discussions

### Mortality

The criterion ‘Must predict mortality’ was rejected. Whilst 8% of voters supported this criterion and 54% felt that it should be included as part of criterion 2, the 70% threshold was not met. Initially, it was highlighted that mortality is a given, so if a mortality-related criterion were to be included, a more appropriate wording might be ‘Associated with an increased risk of mortality’. This would reflect the notion that the incidence of death in a population or cohort with a specific ageing-related pathology, disease or syndrome would be higher than in a pathology-free cohort and/or that ageing-related pathologies shorten life.

However, following extensive discussion, the consortium felt that, on balance, it would be inappropriate to accept a mortality-based criterion. This was on the basis that many ageing-related pathologies or diseases do not have a *direct* impact on risk of death but may have a moderating role, for example hearing loss or osteoarthritis, and that there are many confounding factors involved in the risk of death.

### Inherited/ Mendelian disorders

The original criterion ‘Mendelian disorders are excluded’ had been suggested primarily to exclude diseases or disorders that develop as a result of a highly penetrant, monogenic variant. However, such diseases may have underlying mechanisms that are also observed in ageing-related pathologies, and it could be clinically important to classify and stage such changes, in all contexts. Furthermore, inherited genetic variants, even if they have not yet been identified or are of very low penetrance, can contribute to disease susceptibility, and cannot be ignored in the era of personalised medicine and targeted therapies. The initial criteria would have potentially excluded most progeroid syndromes, which would not be appropriate.

82% of voters disagreed with the inclusion of the suggested criterion, therefore it was rejected.

### Should not be primarily accounted for by an extrinsic carcinogen/ environmental toxin/ infectious agent/ injury

It was agreed not to exclude pathologies, diseases or syndromes that are primarily accounted for by an extrinsic carcinogen, environmental toxin, infectious agent, or injury. This criterion was rejected by 77% of voters on the basis that it is simply not possible to discount the impact of extrinsic or environmental influences on the pathogenesis of many ageing-related pathologies. However, it is acknowledged that there may be specific pathologies, diseases or syndromes that could be excluded by the working groups. For example, the cardiovascular group may decide to include cardiac valve insufficiency as a general ageing-related disease but may exclude insufficiency directly caused by rheumatic fever or acute bacterial endocarditis.

### Frailty

During the Consensus Meeting, there were lengthy discussions surrounding the concept of frailty and whether it should be included, in some way, as a criterion. The term frailty generated debate partly because there are different definitions of frailty, ‘*Frailty is a distinctive health state related to the ageing process in which multiple body systems gradually lose their in-built reserves’* (British Geriatric Society) [8] or ‘*Frailty is a state of increased vulnerability to poor resolution of homeostasis following a stress event, which increases the risk of adverse outcomes including falls, delirium and disability’* (Clegg *et al*.) [9]. Furthermore, different tools are used to measure frailty, for example the Edmonton Frail Scale (EFS) [10] or the Rockwood Clinical Frailty Scale [11] and there is currently no one agreed operational definition.

Frailty is typically accepted as being a syndrome that results in a decreased ability to cope with a stressor [12], and this effectively has been captured in Criterion 2: ‘Should be associated with, or contribute to, functional decline, or an increased susceptibility to functional decline’. In addition to this, not all ageing-related pathologies increase the risk of frailty or are associated with frailty. Therefore, a frailty-based criterion was not accepted.

Only 59 consortium members supported the inclusion of a criterion such as ‘Should be associated with an increased rate of mortality or frailty’ (54 votes) or ‘Should be associated with an increased risk of frailty’ (5 votes).

## Conclusions and future research

The accepted criteria for an ageing-related pathology, disease or syndrome have now been determined and will be used by the ICCARP as the basis for all future classification work. It is important to highlight that the accepted criteria refer to biological and physiological ageing. This project seeks to identify, define and classify pathologies characterised by specific potentially quantifiable changes within cells, tissues and organs, with the acknowledgement that such changes may have a mosaic distribution. The criteria will be re-evaluated in the future to ensure they are still valid in the context of any new research findings.

The next stage of the project is to identify all pathologies that meet the above criteria and to develop proposals for grouping and naming such entities as part of a comprehensive classification system of ageing-related pathologies. This will be done by the system-specific working groups, whilst recognising that there will be several overarching cross-discipline themes that will impact all body systems.

Once the classification phase of the project is complete, this will be followed by defining the criteria for the staging parameters and biomarkers, and to identify these, where possible, for the classified entities. It is recognised that, currently, there may not be clinically validated methods for quantifying many of the pathologies that are classified, and even if there are methods of quantification, there may not be the evidence to determine how severity of a pathology correlates with clinical outcomes. However, it is hoped that, where possible, a systematic approach to identifying and defining biomarkers and staging parameters, for ageing-related pathologies will enable more precise and tailored interventions for ageing populations. Ultimately, such advancements are expected to enhance quality of life and extend healthspan, demonstrating significant personal, societal, economic and healthcare benefits.

## Data Availability

All data produced in the present study are available upon reasonable request to the authors.

## Acknowledgements

This project was supported by a Longevity Impetus Grant from Norn Group.

## International Consortium for the Classification of Ageing-Related Pathologies (ICCARP)

*All listed members are paper co-authors*

